# Association of Time Course of Thrombectomy and Outcomes for Large Acute Ischemic Region: RESCUE-Japan LIMIT Sub-Analysis

**DOI:** 10.1101/2023.03.15.23287338

**Authors:** Hideyuki Ishihara, Takuma Nishimoto, Mototsugu Shimokawa, Fumiaki Oka, Nobuyuki Sakai, Hiroshi Yamagami, Kazunori Toyoda, Yuji Matsumaru, Yasushi Matsumoto, Kazumi Kimura, Reiichi Ishikura, Manabu Inoue, Kazutaka Uchida, Fumihiro Sakakibara, Takeshi Morimoto, Shinichi Yoshimura, the RESCUE Japan LIMIT Investigators

**Author notes:** Corresponding author: Hideyuki Ishihara, Department of Neurosurgery, Yamaguchi University School of Medicine, Minamikogushi 1-1-1, Ube, Yamaguchi, 755-8505, Japan.

## Abstract

**Background:** The effectiveness of endovascular thrombectomy (EVT) has been proven even in patients with large cerebral infarction in early time window. However, the association of the time course with the treatment effect is unknown. The aim of this analysis was to evaluate the influence of the time course from stroke onset to reperfusion on the therapeutic effect of EVT.

**Methods:** The subjects were patients with occlusion of large vessels and sizable strokes on imaging (ASPECTS 3 to 5) in RESCUE-Japan LIMIT (a multicenter, randomized clinical open-label trial of EVT vs. medical care alone). In the current analysis, the clinical and time course characteristics associated with a favorable outcome (modified Rankin Scale (mRS) 0-2 and 0-3 at 90 days) were examined in patients treated with EVT.

**Results:** The analysis included 71 patients (median age, 77 years; median NIHSS score on admission, 21). Occlusion sites were the internal carotid artery (48%), M1 segment of the middle cerebral artery (72%) and tandem lesions (20%). Of these patients, 23 (32%) had mRS 0-3 and 12 (17%) had mRS 0-2 at 90 days. In multivariate analysis, there were independent associations of onset to reperfusion time (OR, 0.991; 95% CI, 0.984-0.999, P = 0.01) and puncture to reperfusion time (OR, 0.952; 95% CI, 0.917-0.988, P < 0.001) with mRS 0-3 at 90 days, and of puncture to reperfusion time (OR, 0.930; 95% CI, 0.872-0.991, P = 0.004) with mRS 0-2 at 90 days.

**Conclusions:** Earlier reperfusion was related to a favorable outcome in patients with acute large vessel occlusion with a large ischemic region. Onset to reperfusion time and especially puncture to reperfusion time were independently associated with a favorable outcome. These results suggest the importance of timing and uninterrupted EVT in this patient population.

## Introduction

Successful reperfusion affects clinical outcomes in patients with acute ischemic stroke (AIS) who have an Alberta Stroke Program Early Computed Tomographic Score (ASPECTS) ≥6, a NIH Stroke Score (NIHSS) ≥6, and causative occlusion of the internal carotid artery (ICA) or proximal MCA (M1) in the early time window.^1^ The Recovery by Endovascular Salvage for Cerebral Ultra-acute Embolism-Japan Large Ischemic Core Trial (RESCUE-Japan LIMIT) also showed efficacy of endovascular thrombectomy (EVT) in patients with a large ischemic core (ASPECTS 3 to 5).^2^ A large ischemic core in the early time window may indicate severe ischemia and the rate of infarct growth may be rapid in such cases. Therefore, effective EVT may require faster treatment, although factors other than time may be more important in such severe cases. In this sub-analysis, we examined the importance of the time factor and the time constraints associated with a favorable outcome.

## Methods

The RESCUE-Japan LIMIT was a multicenter, open-label, randomized clinical trial designed to evaluate the efficacy of EVT with medical care vs. medical care alone in improving clinical outcomes at 90 days after large vessels and sizable strokes seen on imaging (ASPECTS 3 to 5). The study protocol and results have been described elsewhere.^3^ The protocol and consent forms were approved by the institutional review boards at Hyogo College of Medicine and all participating hospitals. Patients or their legally authorized representatives provided written informed consent before enrollment. Patients were eligible if they had acute ischemic stroke with occlusion of the ICA or M1 segment of the MCA on computed tomographic angiography (CTA) or magnetic resonance angiography (MRA), and ASPECTS 3 to 5, as determined on CT or diffusion-weighted MRI; randomization within 6 hours after onset or 6 to 24 hours after the time the patient was last known to be well; no signal change in the initial fluid-attenuated inversion recovery (FLAIR) image, indicating that the infarction was recent; age ≥18, NIHSS ≥6 at admission, and modified Rankin scale (mRS) 0 or 1 before onset of stroke.

RESCUE-Japan LIMIT enrolled 203 patients from November 2018 to September 2021, with follow-up ending in December 2021. The EVT group included 101 patients. Since the accuracy of the time course was particularly important in the current sub-analysis, patients who underwent randomization within 6 to 24 hours after the time they were last known to be well were excluded. Consistent with the main report, the included patients were divided into groups with favorable (mRS 0-3 and 0-2 at 90 days) and poor (mRS 4-6 and 3-6 at 90 days) outcomes. Associations of different intervals from onset to recanalization with favorable outcomes were examined, and the time constraints for a favorable outcome were determined.

### Statistical Analysis

Univariate logistic regression model was used to compare clinical features and different time intervals from onset to recanalization for patients with mRS 0 to 3 vs. 4 to 6 and mRS 0 to 2 vs. 3 to 6 at 90 days. Multivariate logistic regression model was performed to identify independent predictors of two outcomes (mRS 0-3 and mRS 0-2) in cases with ASPECTS 3 to 5 that underwent EVT. Based on previous studies, age, gender, initial NIHSS score, ASPECTS and use of IVT were included for adjusting covariates. The change in adjusted OR every 1 min was analyzed in cases exceeding the time limit. Categorical variables are shown as numbers and percentages, and were analyzed by χ2 test and Fisher exact test, as appropriate. Continuous variables are expressed as median and interquartile range. Some continuous variables were not normally distributed, and these were analyzed by Wilcoxon rank-sum test. All statistical analyses were conducted using JMP 15.0 (SAS Institute Inc, Cary, NC). All reported P values are 2-tailed and P<0.05 was considered to be significant.

## Results

Of 100 patients assigned to EVT in the trial, 71 were included in this sub-analysis, after exclusion of 29 patients with an unknown time of onset (Fig. 1). Of the 71 patients, 23 (32%) had mRS 0-3 and 12 (17%) had mRS 0-2 at 90 days. Baseline characteristics for cases with mRS 0-3 vs. 4-6 and mRS 0-2 vs. 3-6 at 90 days are shown in Table 1.

**Figure 1:**
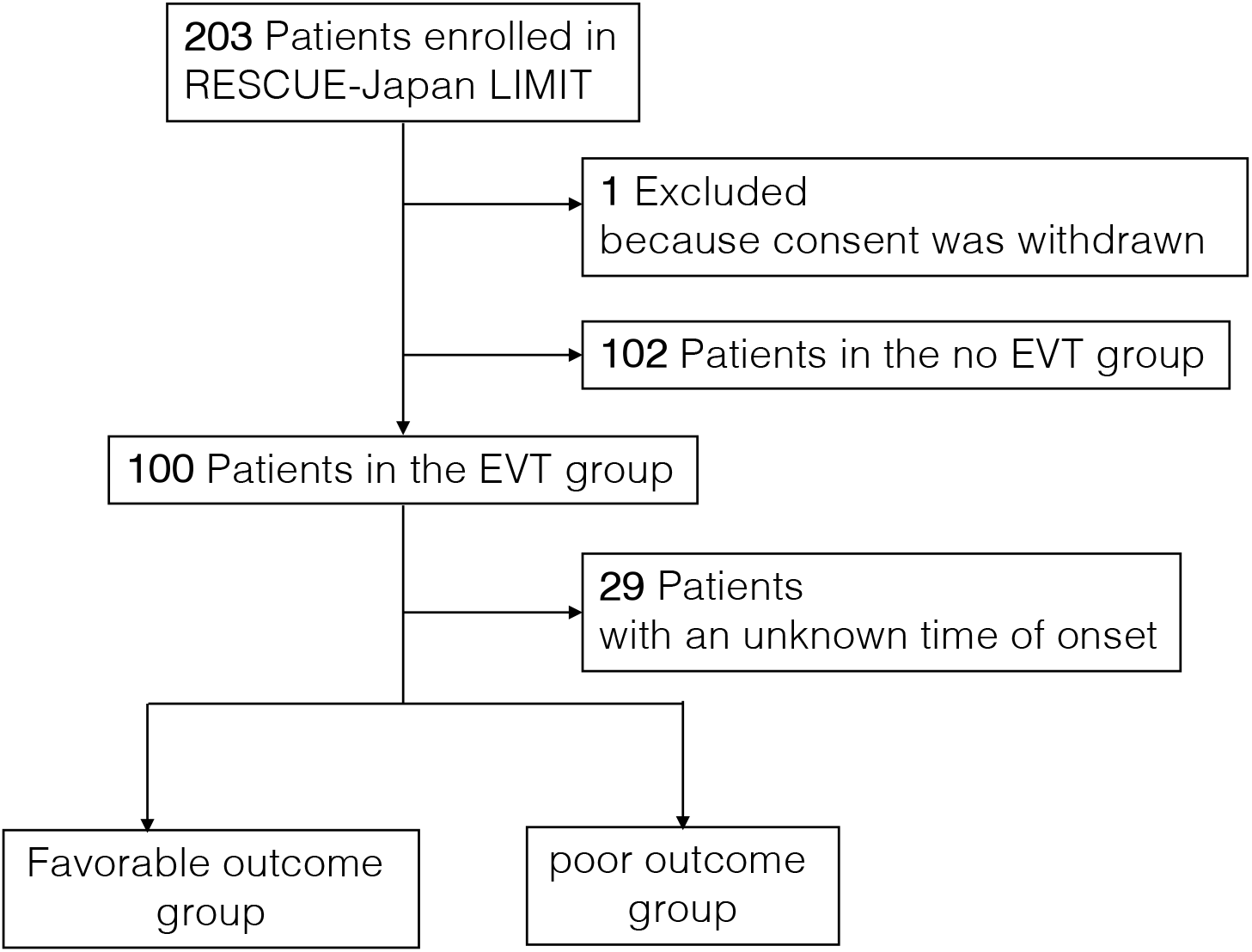
Study Flow Diagram RESCUE-Japan LIMIT, Recovery by Endovascular Salvage for cerebral Ultra-acute Embolism-Japan Large Ischemic Core Trial; EVT, endovascular therapy

**Figure 2:**
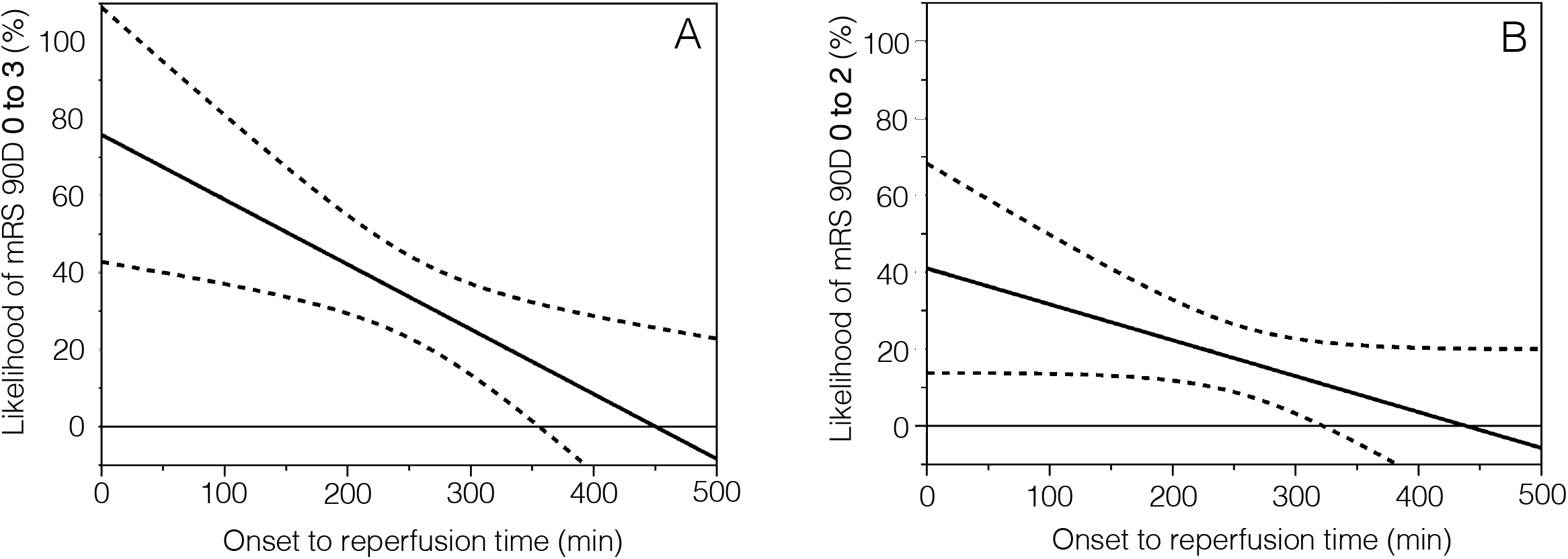
(A) Effect of time on achieving mRS 0-3. (B) Effect of time on achieving mRS 0-2. The association of onset to reperfusion time with likelihood of mRS 0-2 at 90 days is shown. The dashed line indicates the 95% CI.

**Table 1:**
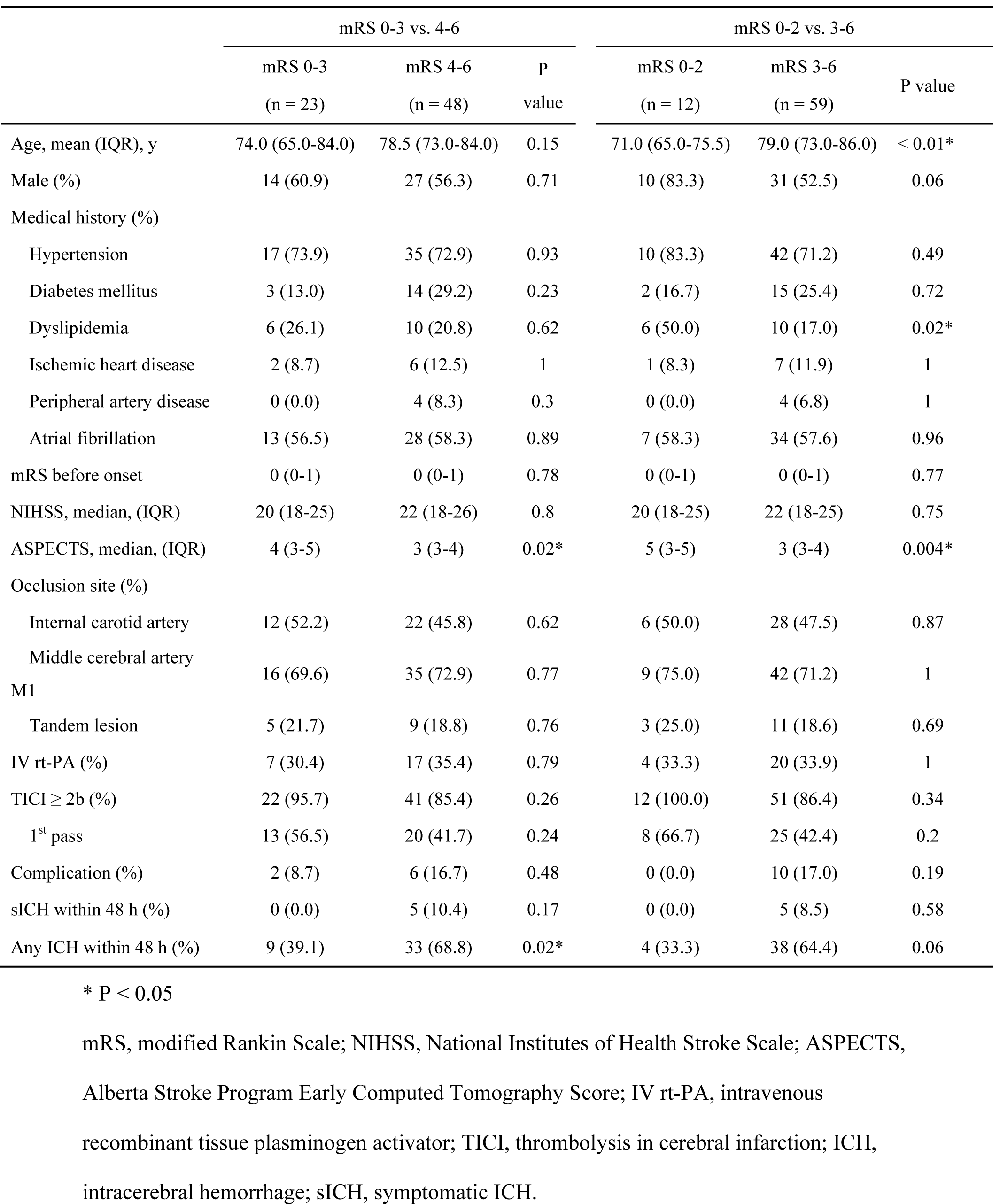
Patient Characteristics

Patients with mRS 0-2 at 90 days were significantly younger than those with mRS 3-6 at 90 days (mean [IQR], 71.0 [65.0-75.5] vs. 79.0 [73.0-86.0] years, P < 0.01); and baseline median ASPECTS was significantly higher in patients with mRS 0-3 vs. 4-6 (4 vs. 3, P = 0.02) and mRS 0-2 vs. 3-6 (5 vs. 3, P = 0.004). Symptomatic ICH within 48 h did not differ significantly for mRS 0-3 vs. 4-6 and mRS 0-2 vs. 3-6, but any ICH within 48 h was significantly less frequent for mRS 0-3 vs. mRS 4-6 at 90 days (9 [39.1%] vs. 33 [68.8%] cases, P = 0.02). There were no significant differences in sex, medical history except for dyslipidemia, mRS before onset, baseline NIHSS, occlusion site, IV rt-PA, reperfusion degree with TICI ≥2b, and frequency of procedural complication between patients with mRS 0-3 and mRS 4-6 at 90 days.

Differences in time courses are shown in Table 2. In univariate logistic regression model comparing patients with mRS 0-3 and mRS 4-6 at 90 days, those with mRS 0-3 had a significantly shorter onset to reperfusion time (median [IQR], 214 [163-250] vs. 291 [205-347] min, P < 0.01), door to reperfusion time (95 [70-136] vs. 118 [89-160] min, P < 0.05), picture to reperfusion time (83 [60-111] vs. 101 [75-162] min, P < 0.05) and puncture to reperfusion time (24 [15-36] vs. 40 [26-61] min, P < 0.001). For mRS 0-2 vs. 3-6 cases, only puncture to reperfusion time was significant (24 [15-34] vs. 37 [25-58] min, P < 0.01) (Table 2).

**Table 2.** Univariate analysis of time course

| Variable | mRS 0-3 vs. 4-6 |  |  | mRS 0-2 vs. 3-6 |  |  |
| --- | --- | --- | --- | --- | --- | --- |
|  | mRS 0-3 | mRS 4-6 | P value | mRS 0-2 | mRS 3-6 | P value |
|  | (n = 23) | (n = 48) |  | (n = 12) | (n = 59) |  |
| Onset to door (min) | 86 (49-160) | 132 (73-212) | 0.09 | 115 (48-153) | 120 (65-212) | 0.19 |
| Onset to picture (min) | 106 (80-170) | 143 (85-214) | 0.11 | 123 (83-139) | 135 (85-215) | 0.31 |
| Onset to randomization (min) | 149 (107-199) | 196 (132-278) | 0.06 | 161 (139-196) | 190 (124-279) | 0.26 |
| Onset to puncture (min) | 165 (135-240) | 223 (156-298) | 0.06 | 185 (156-219) | 215 (140-300) | 0.36 |
| Door to picture (min) | 15 (5-25) | 15 (8-30) | 0.78 | 19 (5-33) | 15 (8-30) | 0.83 |
| Door to puncture (min) | 59 (45-98) | 73 (50-102) | 0.62 | 97 (48-113) | 65 (50-98) | 0.21 |
| Onset to reperfusion (min) | 214 (163-250) | 291 (205-347) | <0.01* | 217 (184-238) | 290 (190-336) | 0.07 |
| Door to reperfusion (min) | 95 (70-136) | 118 (89-160) | <0.05* | 126 (67-151) | 110 (80-160) | 0.67 |
| Picture to reperfusion (min) | 83 (60-111) | 101 (75-162) | <0.05* | 95 (61-118) | 92 (73-155) | 0.51 |
| Puncture to reperfusion (min) | 24 (15-36) | 40 (26-61) | <0.001* | 24 (15-34) | 37 (25-58) | <0.01* |

Values are presented as median (interquartile range) for continuous data.
\* P < 0.05

In multivariate logistic regression model, adjustments were made for age, gender, baseline NIHSS, baseline ASPECTS and IV rt-PA. The adjusted OR for every one minute delay for mRS 0-3 and mRS 0-2 at 90 days are shown in Table 3. Onset to reperfusion time (OR, 0.991; 95% CI, 0.984-0.999, P = 0.01), door to reperfusion time (OR, 0.984; 95% CI, 0.970-0.998, P = 0.01), picture to reperfusion time (OR, 0.986; 95% CI, 0.972-0.999, P = 0.02) and puncture to reperfusion time (OR, 0.952; 95% CI, 0.917-0.988, P < 0.001) were independently associated with mRS 0-3 at 90 days. Only puncture to reperfusion time (OR, 0.930; 95% CI, 0.871-0.991, P < 0.01) was independently associated with mRS 0-2 at 90 days.

**Table 3.** Multivariate analysis of time course

| Variables (/min) | mRS 0-3 vs. 4-6 |  | mRS 0-2 vs. 3-6 |  |
| --- | --- | --- | --- | --- |
|  | Adjusted |  | Adjusted |  |
|  | Odds Ratio<br>(95% CI) | P value | Odds Ratio<br>(95% CI) | P value |
| Onset to door | 0.996 (0.989-1.002) | 0.25 | 0.994 (0.984-1.004) | 0.24 |
| Onset to picture | 0.994 (0.987-1.002) | 0.17 | 0.992 (0.980-1.004) | 0.18 |
| Onset to randomization | 0.994 (0.986-1.002) | 0.11 | 0.993 (0.982-1.004) | 0.19 |
| Onset to puncture | 0.994 (0.986-1.001) | 0.1 | 0.995 (0.985-1.005) | 0.34 |
| Door to picture | 0.999 (0.985-1.013) | 0.88 | 0.999 (0.982-1.0172) | 0.95 |
| Door to puncture | 0.992 (0.974-1.009) | 0.33 | 1.007 (0.986-1.028) | 0.54 |
| Onset to reperfusion | 0.991 (0.984-0.999) | 0.01* | 0.992 (0.982-1.002) | 0.1 |
| Door to reperfusion | 0.984 (0.970-0.998) | 0.01* | 0.993 (0.976-1.009) | 0.36 |
| Picture to reperfusion | 0.986 (0.972-0.999) | 0.02* | 0.993 (0.976-1.009) | 0.37 |
| Puncture to reperfusion | 0.952 (0.917-0.988) | <0.001* | 0.930 (0.871-0.991) | <0.01* |

\* P < 0.05
mRS = modified Rankin Scale
Adjustments were made for age, sex, baseline NIHSS, baseline ASPECTS, use of IV rt-PA

The difference in correlation with mRS 0-3 between onset to reperfusion time and factors used for adjustment was that baseline ASPECTS (OR, 2.067; 95% CI, 1.054-4.052, P = 0.03) and onset to reperfusion time (OR, 0.991; 95% CI, 0.984-0.999, P = 0.01) were significantly associated with mRS 0-3. The difference in correlation with mRS 0-2 between puncture to reperfusion time and factors used for adjustment was that baseline ASPECTS (OR, 3.431; 95% CI, 1.293-9.103, P = 0.007) and puncture to reperfusion time (OR, 0.930; 95% CI, 0.872-0.991, P = 0.004) were significantly associated with mRS 0-2.

Associations of onset to reperfusion times with likelihood of mRS 0-3 and mRS 0-2 at 90 days are shown in Figures 1A and B, respectively. The likelihood of mRS 0-3 at 90 days declined with longer times from symptom onset to reperfusion: 45% at 3 h, and likelihood of mRS 0-3 decreased by 1.7% every 10 min. At an onset to reperfusion time of 6 h, the lower 95% CI for estimated likelihood of mRS 0-3 at 90 days crossed 0 and was no longer significant. Similarly, the likelihood of mRS 0-2 at 90 days declined with longer times from symptom onset to reperfusion: 23% at 3 h, and likelihood of mRS 0-2 decreased by 1% every 10 min. The rate of decline of each likelihood was steeper for mRS 0-3 than for mRS 0-2.

With regard to post-procedural ICH, 5 cases (7%) had symptomatic ICH within 48 h and 42 (59%) had any ICH within 48 h. There were no apparent differences in baseline characteristics of patients with vs. without symptomatic ICH and those with vs. without any ICH (supplemental table 1). In univariate analysis, there was no association between time courses and symptomatic ICH (supplemental table 2) or any ICH within 48 h (Table 4).

**Table 4.** Univariate analysis of time course and any ICH

| Variable | Any ICH +<br>(n = 42) | Any ICH -<br>(n = 29) | P value |
| --- | --- | --- | --- |
| Onset to reperfusion (min) | 257 (194-328) | 227 (168-309) | 0.23 |
| Door to reperfusion (min) | 109 (75-157) | 115 (82-150) | 0.93 |
| Picture to reperfusion (min) | 89 (65-136) | 100 (75-155) | 0.49 |
| Puncture to reperfusion (min) | 36 (23-58) | 34 (21-51) | 0.45 |

ICH, intracerebral hemorrhage

## Discussion

In this study, we report a time-focused sub-analysis of a randomized clinical trial of EVT for patients with acute large stroke in the early time window. This sub-analysis showed that time was an important factor in achieving a favorable outcome, even in cases with a large ischemic region. Among the time intervals, onset to reperfusion time, door to reperfusion time, and puncture to reperfusion were independently associated with mRS 0-3, and puncture to reperfusion time was independently associated with mRS 0-2.

One of the most important goals of treatment for acute large vessel occlusion is to rescue the penumbra region. Since this region is a hypoperfused area that leads to infarction over time, onset to reperfusion time is theoretically the most important time factor. This time has been shown to be associated with outcomes in cases without a large ischemic region in the early time window.^1, 4-8^ Thus, an onset to reperfusion time of 210 min led to an 81% estimated probability of functional independence in patients with occlusion in the proximal anterior intracranial circulation and an absence of large ischemic regions,^7^ and the absolute risk difference for a favorable outcome (mRS 0 to 2) is reduced by 6% to 20% per hour of reperfusion time delay.^6, 7^ Regarding the time limit of onset to reperfusion, the HERMES meta-analysis showed that the efficacy of thrombectomy is no longer significant for achieving mRS 0-2 after 7.3 hours.^1^ The primary endpoint of HERMES was mRS 0-2 at 90 days, while that of Rescue-Japan LIMIT was mRS 0-3 at 90 days. Nevertheless, the present sub-analysis revealed a stricter time constraint of 6 hours in patients with a large ischemic region in the early time window.

There are also reports indicating the importance of other time intervals.^1, 9, 10^ Because the time of stroke onset (last time known well) is often imprecisely determined,^11^ in-hospital time intervals are more strongly associated with outcomes. Each time interval should also contain non-time components relevant to the outcome. Early imaging diagnosis, prompt tPA therapy, and early initiation of endovascular treatment all reflect a well-developed hospital system that includes high diagnostic capabilities and paramedical care. Therefore, reports that door to picture time, door to puncture time,^9, 12, 13^ door to reperfusion time,^1, 14^ picture to reperfusion time^15^ and puncture to reperfusion time^10, 13^ affect clinical outcomes are all reasonable. Furthermore, achieving effective recanalization in one pass (the first pass effect) is strongly associated with favorable clinical outcomes.^16, 17^ The results of this sub-analysis are consistent with these previous reports in patients with no large ischemic regions, and indicate that puncture to reperfusion time is particularly important in patients with large ischemic core regions.

The significance of the RESCUE-Japan LIMIT study was the finding that EVT was effective even in patients with a large ischemic region (ASPECTS 3 to 5) in the early time window. Most cases in the trial were considered to have a malignant profile because of the relationship of ASPECTS with lesion volume.^18, 19^ This profile predicts a poor outcome after thrombolysis due to symptomatic intracerebral hemorrhage (ICH),^20^ and such cases have generally been excluded from clinical trials of EVT. Patients with a malignant profile have been shown to have poor outcomes, even with relatively early recanalization.^19^ In fact, a secondary analysis of RESCUE-Japan LIMIT revealed that EVT was associated with a significantly higher incidence of any ICH within 48 hours in patients with lower ASPECTS.^21^ HERMES also showed that the incidence of symptomatic ICH after recanalization was four times higher in low ASPECTS cases compared with the medical treatment group.^22^ However, the association of time course and ICH after reperfusion in patients with a large ischemic core has not been widely examined.

There is a report that a longer procedural time is associated with symptomatic ICH,^23^ and extending mechanical thrombectomy procedures beyond 60 min increases complications.^24^ Although multiple pass of devices in a procedure results in a longer puncture to reperfusion time,^16, 17^ some meta-analyses suggest achieving reperfusion from the first pass itself is not associated with a reduced symptomatic ICH rate.^25, 26^ The present sub-analysis showed no apparent association between symptomatic or any ICH and time courses. These results might be due to most cases being treated for a short time, and may indicate that time is a critical factor in therapeutic reperfusion with endovascular treatment in patients with large cerebral infarction.

From the perspective of infarction growth velocity, the patients in this study were considered to be fast stroke progressors. Infarction growth rate has shown to be strongly dependent on leptomeningeal collateral flow. The hypoperfusion index ratio obtained from perfusion images and the regional leptomeningeal collateral score are used for assessment of collateral flow, with a hypoperfusion index ratio (T_max_ > 10 volume/ T_max_ > 6 volume) > 0.5 shown to be an indicator of fast infarction growth.^27, 28^ Reperfusion may also prevent infarction growth without increasing the risk of ICH, even in patients with a large volume with T_max_ > 10 s.^29^ These findings are consistent with our results that early recanalization allowed EVT to be effective in patients with a large ischemic region. For such fast stroke progressors, prompt recanalization may be more important than spending time on imaging evaluation. Although controversial, the safety and efficacy of direct transfer to the angiography suite have been shown for patients with suspected large vessel occlusion stroke that may require EVT.^30-32^ Our results indicate that time is critically important for patients with large cerebral infarction in the early time window, and this approach might be particularly effective in these patients.

Following RESCUE-Japan LIMIT study, two randomized trials^33,34^ have shown efficacy of EVT over medical care alone in patients with a large ischemic region. Since both trials included patients within 24 hours after they were last known to be well, the median intervals between the time last known to be well and randomization were about 7 and 9 hours in the two trials. It is difficult to compare the time course of these two trials and those of the present sub-analysis. However, the median puncture to recanalization time was 38 minutes in the SELECT 2 trial.^34^ Thus, based on our results, rapid reperfusion might have contributed to the efficacy of EVT in this trial.

This sub-analysis have several limitations. The sample size was small because only patients who underwent EVT were analyzed and those with an unknown onset time were excluded. However, this allowed analysis using a precise time course and clinical outcomes. The results that onset to reperfusion time and puncture to reperfusion time were especially important in this analysis suggests that these time factors were critically important. The small sample size prevented analysis of each ASPECTS category, but we recently showed in another sub-analysis that EVT is associated with improved outcomes in ASPECTS 4 to 5 cases but not in those with ASPECTS ≤3.^21^ The association of time course and ASPECTS ≤3 is a future issue for consideration. We also note that DWI-MRI was the predominant imaging modality used to evaluate ASPECTS, and DWI-ASPECTS may be approximately one point lower than CT-ASPECTS.^35^ ASPECTS was determined using CT in most previous studies, and thus, a simple comparison with our results for time course should be avoided. Nevertheless, our results are consistent with the trends seen in previous studies.

## Conclusions

Earlier reperfusion was related to effectiveness of EVT in patients with acute large vessel occlusion with a large ischemic region. Onset to reperfusion time and especially puncture to reperfusion time were independently associated with a favorable outcome. These results indicate the importance of timing and uninterrupted endovascular treatment.

## Data Availability

yes

## Appendix

The Recovery by Endovascular Salvage for Cerebral Ultra-acute Embolism-Japan Large Ischemic Core Trial (RESCUE-Japan LIMIT) Investigators’ full names are as follows: Yoshinori Akiyama, Hayato Araki, Takumi Asai, Mikiya Beppu, Yukiko Enomoto, Koichi Haraguchi, Teruyuki Hirano, Hirotoshi Imamura, Ryuzaburo Kanazawa, Naoto Kimura, Norito Kinjo, Shinya Koyama, Naoya Kuwayama, Nobuhisa Matsushita, Masafumi Morimoto, Kenichi Morita, Yoichi Morofuji, Manabu Nagata, Kuniaki Ogasawara, Atsushi Ogata, Tatsuya Ogino, Hiroyuki Ohnishi, Hirotaka Okumura, Tomoya Omae, Takahiro Ota, Takuya Saito, Makoto Sakamoto, Keigo Shigeta, Norihito Shimamura, Seigo Shindo, Fuminori Shimizu, Manabu Shirakawa, Ichiro Suzuki, Masataka Takeuchi, Kanta Tanaka, Kenichi Todo, So Tokunaga, Naoki Toma, Yoshifumi Tsuboi, Wataro Tsuruta, Toshihiro Ueda, Yasushi Ueno, Takehiro Yamada, Yoshitaka Yamada, Nobuaki Yamamoto, Yukako Yazawa, Atsushi Yoshida, Masataka Yoshimura.

## Acknowledgments

We would like to thank all of the Recovery by Endovascular Salvage for Cerebral Ultra-acute Embolism-Japan Large Ischemic Core Trial investigators.

## References

1. Saver JL, Goyal M, van der Lugt A, Menon BK, Majoie CB, Dippel DW, Campbell BC, Nogueira RG, Demchuk AM, Tomasello A, et al. Time to Treatment With Endovascular Thrombectomy and Outcomes From Ischemic Stroke: A Meta-analysis. JAMA. 2016;316:1279–1288. doi: 10.1001/jama.2016.13647

2. Yoshimura S, Sakai N, Yamagami H, Uchida K, Beppu M, Toyoda K, Matsumaru Y, Matsumoto Y, Kimura K, Takeuchi M, et al. Endovascular Therapy for Acute Stroke with a Large Ischemic Region. N Engl J Med. 2022;386:1303–1313. doi: 10.1056/NEJMoa2118191

3. Yoshimura S, Uchida K, Sakai N, Yamagami H, Inoue M, Toyoda K, Matsumaru Y, Matsumoto Y, Kimura K, Ishikura R, et al. Randomized Clinical Trial of Endovascular Therapy for Acute Large Vessel Occlusion with Large Ischemic Core (RESCUE-Japan LIMIT): Rationale and Study Protocol. Neurol Med Chir (Tokyo). 2022;62:156–164. doi: 10.2176/nmc.rc.2021-0311

4. Khatri P, Yeatts SD, Mazighi M, Broderick JP, Liebeskind DS, Demchuk AM, Amarenco P, Carrozzella J, Spilker J, Foster LD, et al. Time to angiographic reperfusion and clinical outcome after acute ischaemic stroke: an analysis of data from the Interventional Management of Stroke (IMS III) phase 3 trial. Lancet Neurol. 2014;13:567–574. doi: 10.1016/S1474-4422(14)70066-3

5. Sheth SA, Jahan R, Gralla J, Pereira VM, Nogueira RG, Levy EI, Zaidat OO, Saver JL, Trialists S-S. Time to endovascular reperfusion and degree of disability in acute stroke. Ann Neurol. 2015;78:584–593. doi: 10.1002/ana.24474

6. Fransen PS, Berkhemer OA, Lingsma HF, Beumer D, van den Berg LA, Yoo AJ, Schonewille WJ, Vos JA, Nederkoorn PJ, Wermer MJ, et al. Time to Reperfusion and Treatment Effect for Acute Ischemic Stroke: A Randomized Clinical Trial. JAMA Neurol. 2016;73:190–196. doi: 10.1001/jamaneurol.2015.3886

7. Goyal M, Jadhav AP, Bonafe A, Diener H, Mendes Pereira V, Levy E, Baxter B, Jovin T, Jahan R, Menon BK, et al. Analysis of Workflow and Time to Treatment and the Effects on Outcome in Endovascular Treatment of Acute Ischemic Stroke: Results from the SWIFT PRIME Randomized Controlled Trial. Radiology. 2016;279:888–897. doi: 10.1148/radiol.2016160204

8. Prabhakaran S, Ruff I, Bernstein RA. Acute stroke intervention: a systematic review. JAMA. 2015;313:1451–1462. doi: 10.1001/jama.2015.3058

9. Bourcier R, Goyal M, Liebeskind DS, Muir KW, Desal H, Siddiqui AH, Dippel DWJ, Majoie CB, van Zwam WH, Jovin TG, et al. Association of Time From Stroke Onset to Groin Puncture With Quality of Reperfusion After Mechanical Thrombectomy: A Meta-analysis of Individual Patient Data From 7 Randomized Clinical Trials. JAMA Neurol. 2019;76:405–411. doi: 10.1001/jamaneurol.2018.4510

10. Enomoto Y, Uchida K, Yamagami H, Imamura H, Ohara N, Sakai N, Tanaka K, Matsumoto Y, Egashira Y, Morimoto T, et al. Impact of Procedure Time on Clinical Outcomes of Patients Who Underwent Endovascular Therapy for Acute Ischemic Stroke. Cerebrovasc Dis. 2021;50:443–449. doi: 10.1159/000515260

11. Spokoyny I, Raman R, Ernstrom K, Kim AJ, Meyer BC, Karanjia NP. Accuracy of First Recorded “Last Known Normal” Times of Stroke Code Patients. J Stroke Cerebrovasc Dis. 2015;24:2467–2473. doi: 10.1016/j.jstrokecerebrovasdis.2015.04.041

12. Jahan R, Saver JL, Schwamm LH, Fonarow GC, Liang L, Matsouaka RA, Xian Y, Holmes DN, Peterson ED, Yavagal D, et al. Association Between Time to Treatment With Endovascular Reperfusion Therapy and Outcomes in Patients With Acute Ischemic Stroke Treated in Clinical Practice. JAMA. 2019;322:252–263. doi: 10.1001/jama.2019.8286

13. Menon BK, Sajobi TT, Zhang Y, Rempel JL, Shuaib A, Thornton J, Williams D, Roy D, Poppe AY, Jovin TG, et al. Analysis of Workflow and Time to Treatment on Thrombectomy Outcome in the Endovascular Treatment for Small Core and Proximal Occlusion Ischemic Stroke (ESCAPE) Randomized, Controlled Trial. Circulation. 2016;133:2279–2286. doi: 10.1161/circulationaha.115.019983

14. Almekhlafi MA, Goyal M, Dippel DWJ, Majoie Cblm, Campbell BCV, Muir KW, Demchuk AM, Bracard S, Guillemin F, Jovin TG, et al. Healthy Life-Year Costs of Treatment Speed From Arrival to Endovascular Thrombectomy in Patients With Ischemic Stroke: A Meta-analysis of Individual Patient Data From 7 Randomized Clinical Trials. JAMA Neurol. 2021;78:709–717. doi: 10.1001/jamaneurol.2021.1055

15. Ribo M, Molina CA, Cobo E, Cerda N, Tomasello A, Quesada H, De Miquel MA, Millan M, Castano C, Urra X, et al. Association Between Time to Reperfusion and Outcome Is Primarily Driven by the Time From Imaging to Reperfusion. Stroke. 2016;47:999–1004. doi: 10.1161/strokeaha.115.011721

16. Zaidat OO, Castonguay AC, Linfante I, Gupta R, Martin CO, Holloway WE, Mueller-Kronast N, English JD, Dabus G, Malisch TW, et al. First Pass Effect: A New Measure for Stroke Thrombectomy Devices. Stroke. 2018;49:660–666. doi: 10.1161/STROKEAHA.117.020315

17. den Hartog SJ, Zaidat O, Roozenbeek B, van Es ACGM, Bruggeman AAE, Emmer BJ, Majoie Cblm, van Zwam WH, van den Wijngaard IR, van Doormaal PJ, et al. Effect of First-Pass Reperfusion on Outcome After Endovascular Treatment for Ischemic Stroke. J Am Heart Assoc. 2021;10:e019988. doi: 10.1161/JAHA.120.019988

18. de Margerie-Mellon C, Turc G, Tisserand M, Naggara O, Calvet D, Legrand L, Meder JF, Mas JL, Baron JC, Oppenheim C. Can DWI-ASPECTS substitute for lesion volume in acute stroke? Stroke. 2013;44:3565–3567. doi: 10.1161/STROKEAHA.113.003047

19. Yoshimoto T, Inoue M, Yamagami H, Fujita K, Tanaka K, Ando D, Sonoda K, Kamogawa N, Koga M, Ihara M, et al. Use of Diffusion-Weighted Imaging-Alberta Stroke Program Early Computed Tomography Score (DWI-ASPECTS) and Ischemic Core Volume to Determine the Malignant Profile in Acute Stroke. J Am Heart Assoc. 2019;8:e012558. doi: 10.1161/JAHA.119.012558

20. Singer OC, Kurre W, Humpich MC, Lorenz MW, Kastrup A, Liebeskind DS, Thomalla G, Fiehler J, Berkefeld J, Neumann-Haefelin T, et al. Risk assessment of symptomatic intracerebral hemorrhage after thrombolysis using DWI-ASPECTS. Stroke. 2009;40:2743–2748. doi: 10.1161/STROKEAHA.109.550111

21. Uchida K, Shindo S, Yoshimura S, Toyoda K, Sakai N, Yamagami H, Matsumaru Y, Matsumoto Y, Kimura K, Ishikura R, et al. Association Between Alberta Stroke Program Early Computed Tomography Score and Efficacy and Safety Outcomes With Endovascular Therapy in Patients With Stroke From Large-Vessel Occlusion: A Secondary Analysis of the Recovery by Endovascular Salvage for Cerebral Ultra-acute Embolism-Japan Large Ischemic Core Trial (RESCUE-Japan LIMIT). JAMA Neurol. 2022;79:1260–1266. doi: 10.1001/jamaneurol.2022.3285

22. Roman LS, Menon BK, Blasco J, Hernandez-Perez M, Davalos A, Majoie Cblm, Campbell BCV, Guillemin F, Lingsma H, Anxionnat R, et al. Imaging features and safety and efficacy of endovascular stroke treatment: a meta-analysis of individual patient-level data. Lancet Neurol. 2018;17:895–904. doi: 10.1016/S1474-4422(18)30242-4

23. Kass-Hout T, Kass-Hout O, Sun CJ, Kass-Hout TA, Nogueira R, Gupta R. Longer procedural times are independently associated with symptomatic intracranial hemorrhage in patients with large vessel occlusion stroke undergoing thrombectomy. J Neurointerv Surg. 2016;8:1217–1220. doi: 10.1136/neurintsurg-2015-012157

24. Spiotta AM, Vargas J, Turner R, Chaudry MI, Battenhouse H, Turk AS. The golden hour of stroke intervention: effect of thrombectomy procedural time in acute ischemic stroke on outcome. J Neurointerv Surg. 2014;6:511–516. doi: 10.1136/neurintsurg-2013-010726

25. Jang KM, Choi HH, Nam TK, Byun JS. Clinical outcomes of first-pass effect after mechanical thrombectomy for acute ischemic stroke: A systematic review and meta-analysis. Clin Neurol Neurosurg. 2021;211:107030. doi: 10.1016/j.clineuro.2021.107030

26. Abbasi M, Liu Y, Fitzgerald S, Mereuta OM, Arturo Larco JL, Rizvi A, Kadirvel R, Savastano L, Brinjikji W, Kallmes DF. Systematic review and meta-analysis of current rates of first pass effect by thrombectomy technique and associations with clinical outcomes. J Neurointerv Surg. 2021;13:212–216. doi: 10.1136/neurintsurg-2020-016869

27. Olivot JM, Mlynash M, Inoue M, Marks MP, Wheeler HM, Kemp S, Straka M, Zaharchuk G, Bammer R, Lansberg MG, et al. Hypoperfusion intensity ratio predicts infarct progression and functional outcome in the DEFUSE 2 Cohort. Stroke. 2014;45:1018–1023. doi: 10.1161/STROKEAHA.113.003857

28. Puhr-Westerheide D, Tiedt S, Rotkopf LT, Herzberg M, Reidler P, Fabritius MP, Kazmierczak PM, Kellert L, Feil K, Thierfelder KM, et al. Clinical and Imaging Parameters Associated With Hyperacute Infarction Growth in Large Vessel Occlusion Stroke. Stroke. 2019;50:2799–2804. doi: 10.1161/STROKEAHA.119.025809

29. Nogueira RG, Haussen DC, Dehkharghani S, Rebello LC, Lima A, Bowen M, Belagaje S, Anderson A, Frankel M. Large Volumes of Critically Hypoperfused Penumbral Tissue Do Not Preclude Good Outcomes After Complete Endovascular Reperfusion: Redefining Malignant Profile. Stroke. 2016;47:94–98. doi: 10.1161/strokeaha.115.011360

30. Requena M, Olive-Gadea M, Muchada M, Hernandez D, Rubiera M, Boned S, Pinana C, Deck M, Garcia-Tornel A, Diaz-Silva H, et al. Direct to Angiography Suite Without Stopping for Computed Tomography Imaging for Patients With Acute Stroke: A Randomized Clinical Trial. JAMA Neurol. 2021;78:1099–1107. doi: 10.1001/jamaneurol.2021.2385

31. Pfaff JAR, Schonenberger S, Herweh C, Ulfert C, Nagel S, Ringleb PA, Bendszus M, Mohlenbruch MA. Direct Transfer to Angio-Suite Versus Computed Tomography-Transit in Patients Receiving Mechanical Thrombectomy: A Randomized Trial. Stroke. 2020;51:2630–2638. doi: 10.1161/STROKEAHA.120.029905

32. Mohammaden MH, Doheim MF, Elfil M, Al-Bayati AR, Pinheiro A, Nguyen TN, Bhatt NR, Haussen DC, Nogueira RG. Direct to Angiosuite Versus Conventional Imaging in Suspected Large Vessel Occlusion: A Systemic Review and Meta-Analysis. Stroke. 2022;53:2478–2487. doi: 10.1161/STROKEAHA.121.038221

33. Huo X, Ma G, Tong X, Zhang X, Pan Y, Nguyen TN, Yuan G, Han H, Chen W, Wei M, et al. Trial of Endovascular Therapy for Acute Ischemic Stroke with Large Infarct. N Engl J Med. 2023. doi: 10.1056/NEJMoa2213379

34. Sarraj A, Hassan AE, Abraham MG, Ortega-Gutierrez S, Kasner SE, Hussain MS, Chen M, Blackburn S, Sitton CW, Churilov L, et al. Trial of Endovascular Thrombectomy for Large Ischemic Strokes. N Engl J Med. 2023. doi: 10.1056/NEJMoa2214403

35. Nezu T, Koga M, Nakagawara J, Shiokawa Y, Yamagami H, Furui E, Kimura K, Hasegawa Y, Okada Y, Okuda S, et al. Early ischemic change on CT versus diffusion-weighted imaging for patients with stroke receiving intravenous recombinant tissue-type plasminogen activator therapy: stroke acute management with urgent risk-factor assessment and improvement (SAMURAI) rt-PA registry. Stroke. 2011;42:2196–2200. doi: 10.1161/STROKEAHA.111.614404

